# Software development for autologous skin substitute production

**DOI:** 10.1101/2021.04.16.21255595

**Authors:** Guillaume Mestrallet

## Abstract

**Objective:** The gold-standard for the management of patients affected by large-surface third-degree burns is autologous skin graft. There is no way to accurately assess burned area size and predict the number of cells and the time necessary to produce a skin graft.

**Materials and Methods:** When burns affect <40% total body surface area (TBSA), meshed skin samples harvested from non-affected donor sites can be used as grafts. In cases corresponding to burns affecting >40% TBSA), the donor site surfaces are insufficient. The alternative grafting strategy uses bioengineered skin substitutes that are generated using the own keratinocytes of the patient after *ex vivo* expansion. We developed a software for severe burn diagnosis and autologous skin substitute production.

**Results:** We developed a software-assisted calculation of the required graft surface and keratinocyte numbers and the time to produce the graft needed, according to patient clinical characteristics. The software also offers assistance to estimate the Baux score, a method that links the severity of burn injuries and the prognosis for the patient.

**Discussion:** Optimal setup of the bioengineering process involved determination of the required graft surface, adjustment of cell quantities, and control of the timing necessary for production. This could be determined and automated by this software. Accordingly, tools to assist the design of personalized protocols will contribute to care quality and cost limitation.

**Conclusion:** This software provides a principle of assisted burned patient diagnose and skin substitute bioengineering process which development may facilitate the design of personalized protocols for skin regenerative cell therapies.

## INTRODUCTION

According to the World Health Organization, burns cause 180,000 deaths a year and more than 30,000,000 cases require hospital treatment in the world in 2018 [1]. Norway is one of the few countries that made public its data on the treatment of burns in 2007. Information from the Norwegian patient registry reveals that a total of 726 patients were admitted to hospitals for acute burns, representing an incidence rate of 15.5 / 100,000 inhabitants for one year [2]. In Norway, the costs of hospital burn treatment in 2007 exceeded 10.5 million euros. The rate of burns requiring hospitalization in children under 5 years was 5.3 times higher or 82.5 / 100,000 inhabitants per year. The average age of all burn patients was 26.9 years, two-thirds of them being men with an average hospital stay of 11.3 days. Fifteen of the patients (2.1%) died of burns in Norwegian hospitals that year. The management of burned patients is still challenging in the 21^st^ century.

The therapeutic arsenal includes split thickness and full thickness skin grafts, when donor sites are available in a sufficient quality and quantity to cover burns [3]. In the context of large-surface burns, the therapeutic option consists in grafting bioengineered skin substitutes generated using patient’s keratinocytes that are massively amplified *ex vivo* to reach required material quantities [4], [5]. Notably, this principle has been combined with viral vector-mediated gene therapy to successfully treat epidermolysis bullosa skin [6], [7]. In skin bioengineering protocols, keratinocytes are extracted from small-size skin biopsies (often 2 or 4 cm^2^) harvested from intact tissue areas, often the groin or armpits (**Figure 1**). Keratinocytes are then amplified for 1 to 3 weeks in bidimensional culture, and then used to generate a three-dimensional sheet [8], [9]. The culture conditions used for keratinocyte amplification originate from the Green’s pioneering work in 1979 [10]. Current processes still use serum and growth-arrested feeder fibroblasts to promote keratinocyte expansion [9].

**Figure 1:**
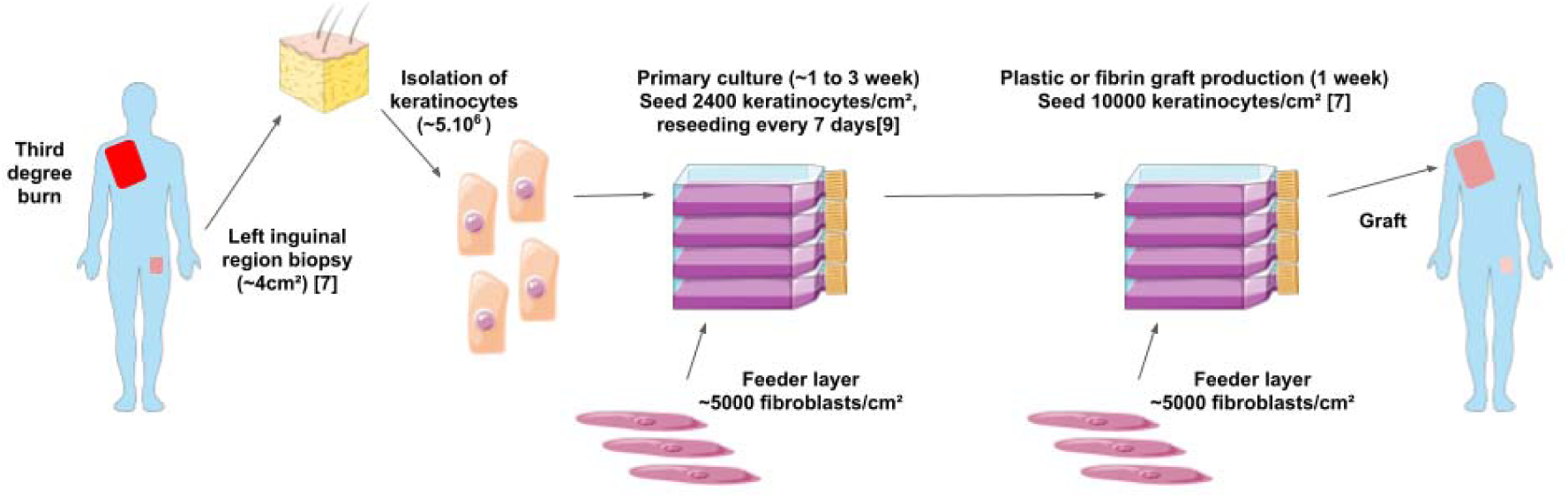
Bioengineering of an epidermal graft.

This technology has proved efficiency to save patient’s lives, as clinical trials have been successfully conducted during the last three decades [8]. For example, one of these included 63 patients and reported a transplant take rate of 65%, with a patient survival rate of 84% after 7 days. The reported infection rate was 4.3%, and 11% of the patients had received a second transplantation procedure [11]. A second one included 88 patients and reported a transplant take rate of 72.7%, with a patient survival rate of 91% [12]. Recently, ten pediatric patients with skin defects were included in a clinical study[13]. Skin grafts of 49 cm were bioengineered using autologous keratinocytes and fibroblasts isolated from a 4cm^2^ patient’s skin biopsy and then incorporated in a collagen hydrogel. In this phase 1 study, safe coverage of skin defects was achieved.

The clinician calculates the Total Burn Surface Area (TBSA) in the second and third degree. The percentage of the total burned surface of the body to which each zone corresponds is calculated according to age, with the rule of Wallace or that of Lund and Browder [15]–[17]. Software development will automate and make more rigorous these calculations, thus allowing a better diagnosis. This work is a proof of principle established from data published in the literature [7], [9].

Keratinocytes are obtained from 2 to 4 cm^2^ skin biopsies from healthy areas that heal well and have little exposure, such as the groin and armpits [6], [8]. Keratinocytes are seeded at 2400 keratinocytes/cm^2^ and preamplified for 1 to 3 weeks in sheets [9].Then, keratinocytes are seeded at 10000 keratinocytes/cm^2^, amplified and transferred onto a sterile gauze that will then be deposited on the area to be treated after 7 more days [7]. Keratinocytes are amplified with a feeder layer of fibroblasts, in serum containing medium.

## METHODS

The software only allows clinicians registered with the authors to have access to medical data, in order to avoid misuse of the data. The software runs autonomously on a computer or on a mobile phone to protect patient confidentiality. Communication between the database and users was done via the Hypertext Transfer Protocol (HTTP) and an Application Programming Interface (API) specific to this software. The protection of personal data, with the new European GDPR legislation, aims to protect patient data. If requested, it may be possible to store this data on the hospital database in connection with the patient’s computerized medical record [23]. The patient information is also anonymised with a medical identifier. Once connected, the clinician can either diagnose a new patient or access the diagnosis of a registered patient by indicating their medical identifier. In **Table 1** are referenced the questions allowing to diagnose the patient via the form.

**Table 1:** Patient information collected by form.

| **Table 1: Patient information collected by form** |
| --- |
| -What is the patient's first name? |
| -What is the patient's name? |
| -What is the date of birth? |
| -What is the weight of the patient (in kg)? |
| -What is the patient height (in cm)? |
| -Is there an inhalation injury (yes/no)? |
| -Indicate the percentage (between 0 and 100) of the head and neck that is affected in the 2<sup>nd</sup> degree. |
| -Indicate the percentage (between 0 and 100) of the head and neck that is affected in the 3<sup>rd</sup> degree. |
| -Indicate the percentage (between 0 and 100) of the anterior face of the trunk which is affected in the 2<sup>nd</sup> degree. |
| -Indicate the percentage (between 0 and 100) of the anterior surface of the trunk which is affected in the 3<sup>rd</sup> degree. |
| -Indicate the percentage (between 0 and 100) of the posterior surface of the trunk which is affected in the 2<sup>nd</sup> degree. |
| -Indicate the percentage (between 0 and 100) of the posterior surface of the trunk which is affected in the 3<sup>rd</sup> degree. |
| -Indicate the percentage (between 0 and 100) of the upper limbs that is affected in the 2<sup>nd</sup> degree. |
| -Indicate the percentage (between 0 and 100) of the upper limbs that is affected in the 3<sup>rd</sup> degree. |
| -Indicate the percentage (between 0 and 100) of the lower limbs that is affected in the 2<sup>nd</sup> degree. |
| -Indicate the percentage (between 0 and 100) of the lower limbs that is affected in the 3<sup>rd</sup> degree. |
| -Indicate the percentage (between 0 and 100) of the external genital organs that is affected in the 2<sup>nd</sup> degree. |
| -Indicate the percentage (between 0 and 100) of the external genitalia that is affected in the 3<sup>rd</sup> degree. |

The clinician calculates the Total Burn Surface Area (TBSA) in the second and third degree. In accordance with current clinical standards, these areas are the head and neck, the lower limbs, the upper limbs, the external genitalia, the posterior surface of the trunk and the anterior surface of the trunk. However, the percentage of the total area of the body that these different areas represent varies depending on the age of the patient. Informing the patient’s age therefore allows these different areas to be assigned an appropriate value. Each value represents a percentage of the total body surface area based on the age of the patient. The calculation of the total area of the patient’s body from his weight and height is done using the Du Bois formula [14].

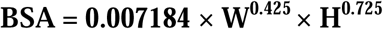

With BSA = Body Surface Area in m^2^, W = weight in kg and H = height in cm.

Once the form has been completed, the software calculates the variables in **Table 2** and displays the patient’s diagnosis.

**Table 2:** Diagnostic variables calculated by the algorithm.

| **Table 2: Diagnostic variables calculated by the algorithm** |
| --- |
| -total area of the patient burned in the 2<sup>nd</sup> and 3<sup>rd</sup> degree ( % of total body area) |
| -total area of the patient burned in the 2<sup>nd</sup> degree ( % of total body area) |
| -total area of the patient burned in the 3<sup>rd</sup> degree to be grafted on as soon as possible ( % of total body area) |
| -Graft surface to prepare (in cm<sup>2</sup>) |
| -Surface of the biopsy to be taken to prepare the autologous graft (in cm<sup>2</sup>) |
| -Duration of amplification in culture to obtain the graft (in days) |
| -Graft preparation protocol |
| -Baux score |
| -Probability of death (%) |

The percentage of the total surface of the body to which each zone corresponds is calculated as in **Table 3**, according to age, with the rule of Wallace or that of Lund and Browder [15]– [17].

**Table 3:** Assessment of the burned area as a function of age.

| **Table 3: Assessment of the burned area as a function of age** |  |  |  |  |
| --- | --- | --- | --- | --- |
|  | Age (years) |  |  |  |
|  | 0-4 | 5-9 | 10-15 | >15 |
| Head and neck | 21% | 15% | 12% | 9% |
| Anterior face of the trunk | 13% | 13% | 13% | 18% |
| Posterior surface of the trunk | 18% | 18% | 18% | 18% |
| Upper limbs | 19% | 19% | 19% | 18% |
| Lower limbs | 28% | 34% | 37% | 36% |
| External genital organs | 1% | 1% | 1% | 1% |

The size of the graft to be prepared is calculated by summing the burned areas in the third degree. The size of the skin biopsy to be taken and the graft preparation protocol is determined by previous clinical observations [7]. Keratinocytes have a high proliferation rate and grow through a total of 20 to 50 generations depending on the age of human donor, 20 generations correspond to an increase in cell mass of approximately 106-fold [18]. From a single cm^2^ of biopsy of healthy tissue, it is possible to extract 1,000,000 to 4,000,000 keratinocytes depending on the age and anatomic site [7]. The choice was made to fix the maximum size of the sample at 4cm^2^ to avoid unduly altering the unaffected areas of the patient.

Thus, keratinocytes are obtained from 2 to 4 cm^2^ skin biopsies from healthy areas that heal well and have little exposure, such as the groin and armpits [6], [8]. Keratinocytes are amplified for 1 to 3 weeks with successive passages every 7 days. Keratinocytes are amplified with a feeder layer of fibroblasts, in serum containing medium. There is a period of amplification of the cells ex vivo, followed by a period of preparation of the graft, with different seeding densities. The seeding densities for these two periods are referenced in **Figure 1** and are taken from previous studies [7]. In this last publication, 3×10^7^ keratinocytes provide 2880cm^2^ in 7 days with fibrin. It is therefore necessary to inoculate 10417 keratinocytes for 7 days on fibrin in order to have 1cm^2^ of skin. This figure is here rounded to 10,000. For the preamplification phase, keratinocytes are thawed and plated at 2,400 cells per cm2 [9]. The production time is calculated based on the size of the burned area. If the number of cells required for seeding for graft production is less than or equal to the number of cells obtained from the biopsy, the production time is 7 days. If the number of cells necessary for the seeding for graft production is between the number of cells obtained from the biopsy and the number of cells that can be obtained at most after 7 days of amplification, the production time is 14 days. If it is higher, the production time is 21 days.

The number of cells needed to prepare the graft and the size of the biopsy to be taken to obtain them depends on the preparation time of the graft chosen. We define two new formulas to calculate them.

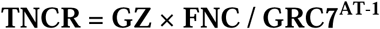

With TNCR = total number of cells required for the first amplification during the first 7 days, GZ = graft size (cm^2^), FNC = Final number of cells to seed per cm^2^ 7 days before grafting. At this stage, 7 days before the transplant, it goes from cell amplification to graft production. GRC7 = growth rate of cells in 7 days (average number of cells obtained from a single cell), AT = amplification time (weeks).

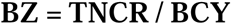

With BZ = biopsy size (cm^2^), BCY = biopsy cell yield (number of cells extracted from a biopsy cm^2^). Here FNC is equal to 10,000 cells/cm^2^, GRC7 of keratinocytes is equal to 17 cells and BCY is equal to 1,250,000 cells/cm^2^ [7].

Finally, the revised Baux score for mortality prediction in burns patients has been adopted [19].

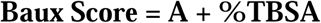

With A = Age in years, %TBSA = Percentage of Total Body Surface Area burned (between 0 and 100).

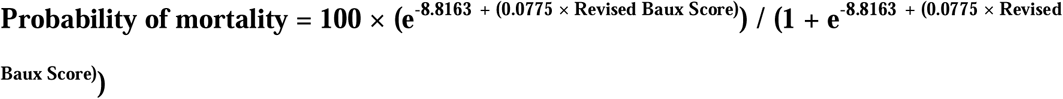

With Revised Baux Score = Baux Score +17 if there is an inhalation injury.

## RESULTS

As each laboratory or production company uses its own calculation parameters to produce a graft, it is possible for them to indicate them on their software interface so that the calculations adapt to their production methods. The results of **Figure 2** are obtained by fixing the amplification and production parameters according to previous studies (see introduction and methods) and by varying the amplification time between 7, 14 and 21 days. Here we take the values drawn from the work of the De Luca team [7] and Percy hospital [9]. AT varies from 1 to 3 weeks. As can be seen, an amplification of a single week can produce a graft of sufficient size only if the surface to be grafted is very small (< 500cm^2^). Indeed, it would otherwise be necessary to carry out a too large biopsy, incompatible with the patient’s survival. Likewise, an amplification of 2 weeks only makes it possible to produce a graft of sufficient size if the surface to be grafted is not too large in size (< 8,500cm^2^). On the other hand, an amplification of 3 weeks makes it possible to produce a graft of a size sufficient to cover an entirely burned patient (∼ 20,000cm^2^) from a biopsy of less than one cm^2^. However, waiting so long may be incompatible with patient survival.

**Figure 2:**
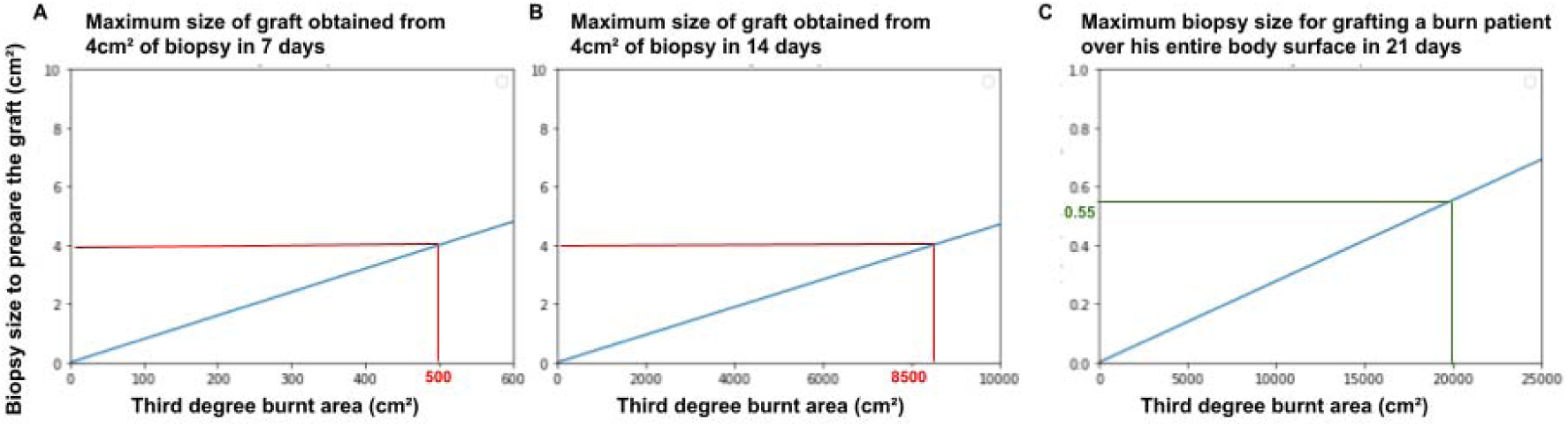
Calculation of the biopsy size to prepare the graft, according to the amplification time.

Calculation of the biopsy size (cm^2^) to produce a graft of sufficient size, according to the duration of amplification (weeks). A : 1 week, B : 2 weeks, C : 3 weeks.

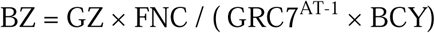

With GZ = graft size (cm^2^), FNC = Final number of cells to seed per cm^2^ 7 days before grafting, GRC7 = growth rate of cells in 7 days (average number of cells obtained from a single cell), AT = amplification time (weeks), BZ = biopsy size (cm^2^), BCY = biopsy cell yield (number of cells extracted from a biopsy cm^2^).

The software makes it possible to determine graft preparation scenarios according to the area burned in the third degree and therefore the size of the graft to be prepared. They are presented in **Table 4**. These scenarios are characterized by the size of the graft, the duration of its preparation and the size of the biopsy of healthy tissue to be taken from the patient. We chose to limit the size of the biopsy to 4cm^2^ to avoid unduly altering the unaffected areas of the patient [7].

**Table 4:** Graft preparation scenarios calculated by the software.

| **Table 4 : Graft preparation scenarios calculated by the software** |  |  |  |
| --- | --- | --- | --- |
| Third degree burned area (cm<sup>2</sup>) | $x < 500$ | $500 < x < 8500$ | $8500 < x < 144500$ |
| Graft preparation time (days) | 12 | 21 | 28 |
| Size of biopsy to be taken (cm<sup>2</sup>) | < 4 | 4 | 4 |

In the case of a third degree burned surface of less than 500cm^2^, it is not necessary to take more than 4cm^2^ of tissue and the graft can be prepared in 12 days. The keratinocytes must be extracted from the biopsy and amplified in co-culture with a feeder layer of fibroblasts according to current clinical protocols. In the case of a third degree burned area between 500cm^2^ and 8,500cm^2^, 4cm^2^ of tissue must be removed and the graft can be prepared in ∼ 20 days. It is necessary to extract the keratinocytes from the biopsy and to amplify them in co-culture with a feeder layer of fibroblasts according to current clinical protocols for 12 days, trypsinize the cells from the initial cell culture flasks and re-amplify them 7 days after seeding them a the appropriate density. In the case of a third degree burned surface greater than 8,500cm^2^, the same procedure must be followed, adding another 7 days of amplification. The total body surface of an adult human is approximately 2m^2^ (20,000cm^2^) so the preparation of the graft does not require more than 28 days in the worst case.

The software considers these different scenarios to select the one that is the most suitable. In **Figure 3**, there is an example of the data collection of a fictitious patient carried out by a clinician as described in the methods section. These data are processed and saved in the database to allow the clinician to find them later if necessary.

**Figure 3:**
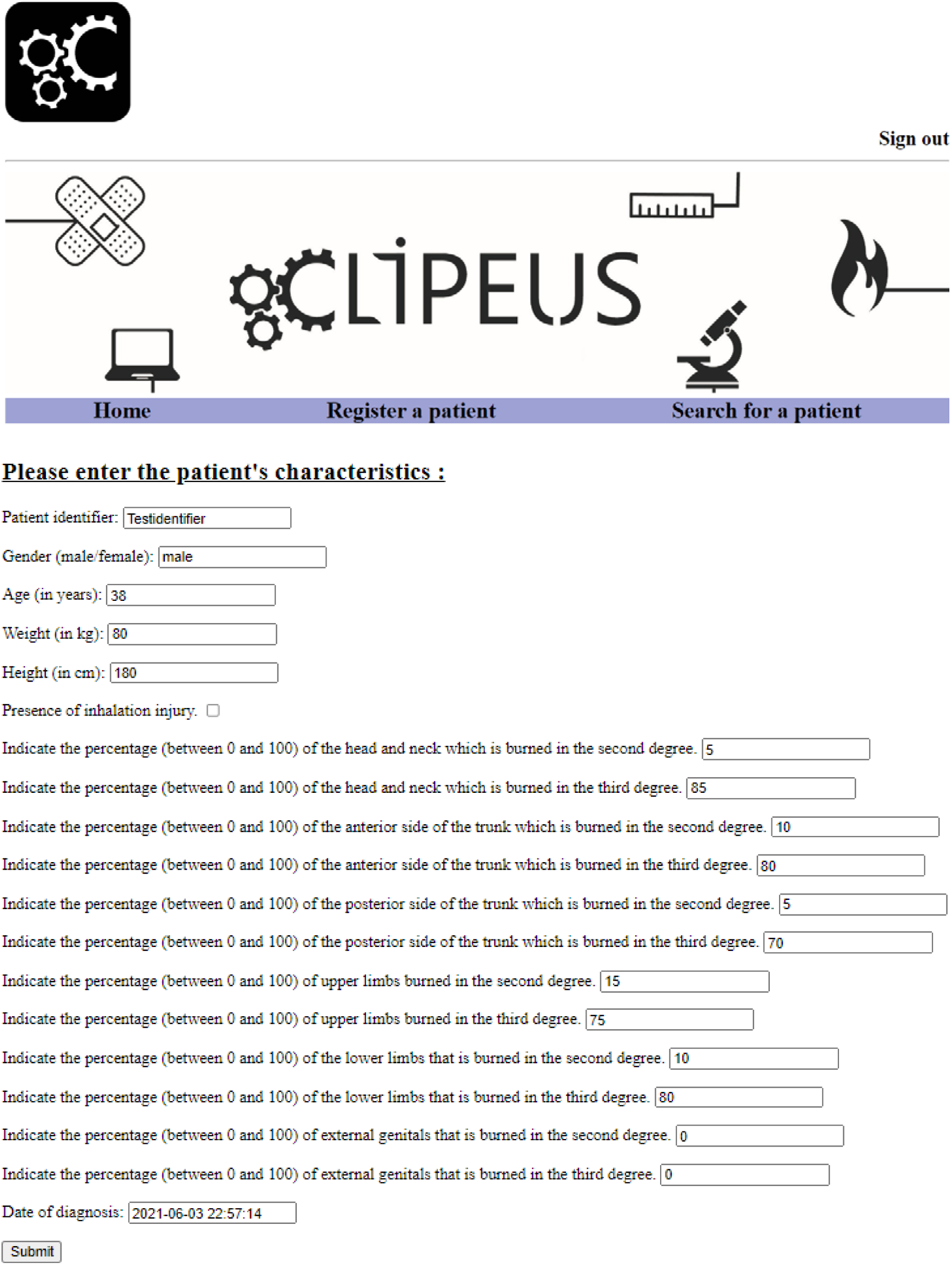
Burned patient data collection form.

Example of the data collection from a fictitious patient by a clinician as described in the methods section.

In **Figure 4**, there is an example of a fictitious patient diagnosis by the software as described in the methods section. The fictitious patient is therefore in the situation corresponding to the second scenario in **Table 4**. His probability of mortality is 66% but it takes 21 days to produce the graft from a 4cm^2^ biopsy. The size of the graft to be produced corresponds to the third degree burned area, which is 15363 cm^2^ here. The graft production protocol is then described with the number of cells required for each step.

**Figure 4:**
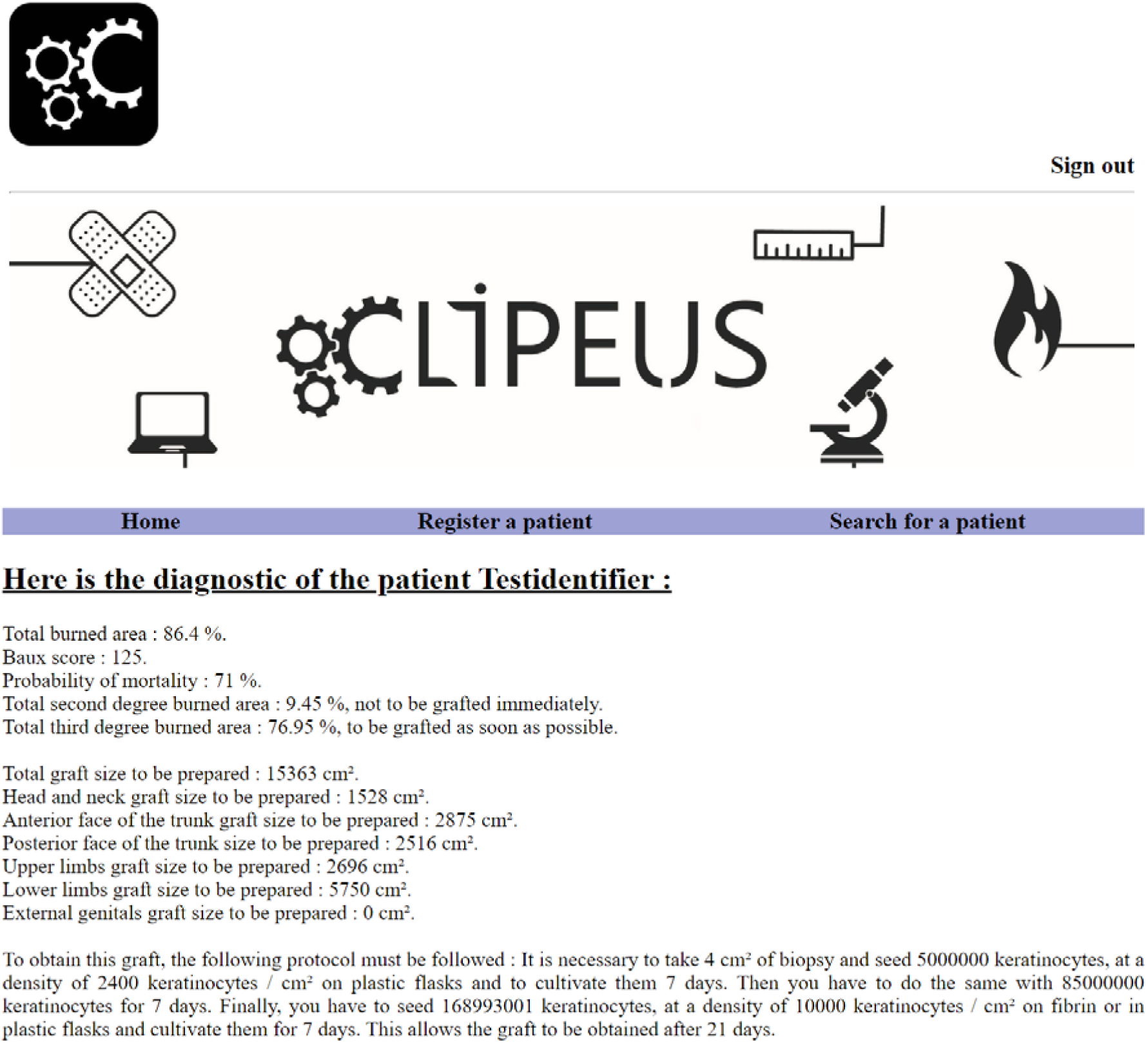
Diagnosis of the burned patient.

Example of the diagnosis of a fictitious patient calculated by the software as described in the methods section.

## DISCUSSION

Few quantitative methods are reported in the literature to adjust the cell quantities necessary in view of skin substitute production according to specific patient characteristics. No automation and quantification allowing diagnosis are proposed. One rare quantitative approach using Matlab computing language is tensor decomposition for color image segmentation of burn wounds [20]. The aim is to use a new automated segmentation of the images in order to determinate the burn area. An issue is to investigate the segmentation of burn areas on 3D images to include curves and depth to further improve the segmentation accuracy [22]. The software developed here is complementary to this “Burn Case 3D” work, because it allows to calculate the number of cells and the duration necessary for the graft production.

The software developed here enables segmentation of the burned areas to be treated as a priority. By automating the calculation of the area to be transplanted in the patient, the duration of preparation of the graft and the quantity of cells required, it allows the clinician to plan the treatment more reliably. By defining two new formulas to calculate the number of cells required to prepare the graft and consequently the size of the associated biopsy, it makes it possible to prepare a graft in a more precise time with the least possible materials and costs. The software proposes three different bioengineering scenarios in two, three or four weeks (**Table 4**), which can help the clinician to predict the patient’s treatment time and the length of hospital stay.

According to the data from the 2007 Norwegian patient registry, the average total cost of treating a patient with hospital burns is around 14 463 €, which corresponds to 1 280 € per night [2]. The use of this software can help to better calculate the time spent by the patient in the hospital. Its use could therefore save 1 280 € per night and per patient in the event of overestimation of the duration of graft preparation.

In case of a deep burn in the hand (80% of cases), there are therapeutic alternatives to excision-grafting [21]. These are artificial dermis (Alloderm, Matriderm, Integra) and flaps. In the presence of immediately usable donor areas, the combination Matriderm / full thin skin graft is recommended. However, these techniques are expensive. The Integra® (artificial dermis) costs between 5 and 5.5 € per cm^2^, the Matriderm®, between 4 and 5 € per cm^2^, while the Alloderm is even more expensive (32 € 50 per cm^2^). Moreover, there is always a need to best estimate the size of the graft, so the program developed here is also useful in these cases. Take the case of an adult with third degree burns on 10% of the upper limbs. His total body surface area is approximately 2 m^2^. An overestimation of 10% of the size of the burn, and therefore of the size of the commercial graft to be ordered, can lead to an additional cost of 36 to 360 €.

Today, although the technology for producing autografts is not new, there is no way to accurately assess burned areas and predict the number of cells necessary to produce the graft. Developing a skin substitute with the help of softwares to accurately estimate the needs would optimize the whole process. The new software presented here is the first one to better automate the evaluation of burn size and that of the graft to be prepared. It is also the first to calculate the number of cells and the duration necessary for the graft production, as well as the size of the biopsy of healthy tissue to be taken, the Baux score and the probability of mortality. The limitation of this software is the differences in cell quality for skin regeneration between various donors, due to interindividual variations, well known in the cellular biology scientist’s community. This software therefore brings progress towards more precise determination of the treatment conditions of severely burned patients.

## Data Availability

All data are included in the paper.

https://gitlab.com/gmestr/skin

## ACKNOWLEDGEMENTS

The author thanks Alexandre Pierga for the visual elements of the software, Clément Hénin and Louis-Gabriel Mognetti for their contribution to technical aspects of the software, and Dr Lellouch for clinic-related discussions. This research did not receive any specific grant from funding agencies in the public, commercial, or not-for-profit sectors.

## ABBREVIATIONS

A: Age
API: Application Programming Interface
AT: amplification time
BCY: biopsy cell yield (number of cells extracted from a biopsy cm^2^)
BSA: Body Surface Area
BZ: Biopsy Size
GRC7: Growth Rate of Cells in 7 days (average number of cells obtained from a single cell)
GZ: Graft Size
H: Height
HTTP: Hypertext Transfer Protocol
FNC: Final number of cells to seed per cm^2^ 7 days before grafting
STSG: Split Thickness Skin Graft
TBSA: Total Body Surface Area
TNCR: Total Number of Cells Required
WHO: World Health Organization
W: Weight
%TBSA: Percentage of Total Body Surface Area burned

